# Accuracy of preferred language data in a multi-hospital electronic health record in Toronto, Canada

**DOI:** 10.1101/2025.02.27.25323003

**Authors:** Camron D Ford, Thomas Bodley, Martin Betts, Rob Fowler, Alexis Gordon, Michele James, Shail Rawal, Christina Reppas-Rindlisbacher, Paul Tam, George Tomlinson, Christopher J Yarnell

## Abstract

**Background:** Accurate preferred language data is a prerequisite for providing high-quality care. We investigated the accuracy of preferred language data in the electronic health record (EHR) of a large community hospital network in Toronto, Canada.

**Methods:** We conducted a point-prevalence audit of patients admitted to intensive care, internal medicine, and nephrology services at three hospitals. We asked each patient “What is your preferred language for health care communication?” and reported on agreement (with 95% confidence intervals [CI]) between interview-based and EHR-based preferred language. We used Bayesian multilevel logistic regression to analyze the association between patient factors and the accuracy of the EHR for patients who preferred a non-English language.

**Results:** Between June 17, 2024, and July 19, 2024, we interviewed 323 patients, of whom 124 (38%) preferred a non-English language. Median age was 77 years and 46% were female. EHR accuracy was 86% for all patients. The probability of the EHR correctly identifying a patient with non-English preferred language (sensitivity) was 69% (CI 60 to 77), specificity was 97% (CI 94 to 99), positive predictive value was 95% (CI 88 to 98), and negative predictive value was 83% (CI 79 to 87). There were 26 different non-English preferred languages, most commonly Cantonese (27%) and Tamil (14%). Accuracy was better for patients who were female or older, and varied by hospital and medical service.

**Conclusions:** In this multi-hospital point-prevalence audit, the EHR accurately captured language preference for 86% of all patients and 69% of patients who preferred a non-English language.

## Introduction

Patients who do not speak the language in which their care is provided are at higher risk of harm and poor outcomes. Canadian data shows an 18-30% higher risk of harm for hospital inpatients who do not speak English, compared to inpatients who do speak English.(1,2) Such patients are common in Canada, where 2021 census data showed 690,000 people who did not speak either official language. In the United States, 5,720,834 (4.4%) households were classified as “limited English-speaking” in the 2023 American Community Survey.(3) Neither statistic includes people who may speak basic English, but would prefer to discuss complicated medical decisions in another language. Language concordant care, where a patient receives care in their preferred language for healthcare communication, can help address potential disparities.

Language concordance, via shared language proficiency or interpretation, is essential for effective care. It helps clinicians to understand patient symptoms and values in order to make informed recommendations, and it helps patients to understand their situation and options, in order to make informed decisions.(1,2) Ensuring access to communication, independent of a patient’s preferred language, is a pillar of equitable care.(4,5) Given the importance of language-concordant care, clinicians and healthcare systems need accurate data regarding their patients’ preferred language.

The accuracy of preferred language data in electronic health records (EHRs) varies. A study conducted in 2020 in Toronto, Ontario, found sensitivities of 81% and 12% at two academic hospital sites.(6) Another study conducted in an outpatient setting found that the EHR accurately identified preferred language in two-thirds of Spanish-speaking patients.(7) Previous Canadian evaluations involved only academic hospitals and uncommon EHRs.(8) To inform quality improvement and research efforts aimed at improving language concordant care, we evaluated the accuracy of the preferred language field in a commonly used EHR (Epic Systems) at three large community hospitals.

## Methods

We performed a prospective point-prevalence audit of inpatients at three community hospitals within the same health network in Toronto, Canada, to evaluate the accuracy of the preferred language field in the EHR. We used an approach similar to prior work.(6) The audit was deemed quality improvement by the Scarborough Health Network research ethics board which granted a waiver of the requirement for informed consent (PQI-24-002).

One investigator (CF) prospectively interviewed patients admitted to the intensive care unit, internal medicine ward, and nephrology ward across the three hospitals. On each interview day, they selected a particular geographic ward of the hospital containing approximately 30-50 patients and proceeded sequentially from room to room. After introducing themselves and the project to the patient, they asked one question: “What is your preferred language for health care communication?”(6) We recorded the response as “English” or “Not English.” In the case where the preferred language was not English, we recorded the preferred language.

If the patient was unavailable at the time of interview, or a response could not be elicited, a substitute decision maker (SDM) was contacted and their response to the question was recorded. If an SDM was unable to be reached upon the first attempt, a second attempt was made. Upon an unsuccessful second attempt, the patient was omitted from the audit.

We recorded interview-based preferred language and EHR-based preferred language in a REDCap database.(9) We also collected age, sex, hospital of admission, most responsible service, and length of stay. In the primary analysis, we calculated the sensitivity, specificity, positive predictive value, and negative predictive value of using EHR-based non-English preferred language to identify true non-English language preference.

The secondary analysis focused on identifying factors associated with correctly identifying non-English preferred language. We did this by modeling the probability of an EHR-based non-English preference among patients who had an interview-based non-English preference using Bayesian logistic regression. Fixed predictors were age, sex, hospital, length-of-stay, and inpatient service; preferred language was a random effect, to allow for the incorporation of languages with few respondents without causing overfitting.(10) We modeled age in decades to make its odds ratio more interpretable. We modeled the logarithm (base 2) of length-of-stay to aid in model convergence. We reported ORs, including the median OR which measures the extent of variability between clusters (languages).(11,12) We used a Bayesian approach because when clusters are small, frequentist multilevel models may have difficulties with convergence.(10)

We reported uncertainty using 95% confidence intervals (CI) in the primary analysis, and 95% credible intervals (CrI) in the secondary analysis. Models were fit in R using the brms package.(13,14)

## Results

Between June 17, 2024, and July 19, 2024, we interviewed 323 patients across 3 hospitals (Table 1). The number of included patients per hospital ranged from 85 to 108. The median age was 77 years (Interquartile range [IQR]: 65-85) and 46% of patients were female. The most common responsible service was internal medicine. For 132 patients (41 %), we obtained preferred language information from the substitute decision-maker.

**Table 1:** Characteristics of Patients List of audit sample characteristics grouped by hospital site. ICU = intensive care unit. IQR = interquartile range.

| Patient Characteristic | Hospital Site |  |  | Total |
| --- | --- | --- | --- | --- |
|  | 1<br>n=130 | 2<br>n=85 | 3<br>n=108 |  |
| **Female (%)** | 56 (43%) | 41 (58%) | 53 (49%) | 150 (46%) |
| **ICU (%)** | 31 (24%) | 22 (26%) | 6 (6%) | 59 (18%) |
| **Nephrology (%)** | 26 (20%) | 0 | 0 | 26 (8%) |
| **Medicine (%)** | 73 (56%) | 63 (74%) | 102 (94%) | 238 (74%) |
| **Median age, years (IQR)** | 73 (63 – 83) | 75 (63-82) | 82 (71-90) | 77 (65-85) |
| **Median length of hospital stay, days (IQR)** | 10 (4 – 25) | 7 (4 – 14) | 8 (4-24) | 9 (4 – 23) |

The number of patients who preferred a non-English language was 93 (29%) by EHR, and 124 (38%) by interview (Table 2, absolute difference of 9.6%, CI 2.4 to 16.8, p < 0.01). There were 26 different languages represented among the interview-based non-English preferred languages. The most common non-English preferred languages were Cantonese (26.6%), Tamil (13.7%), and Italian (8.1%).

**Table 2:**
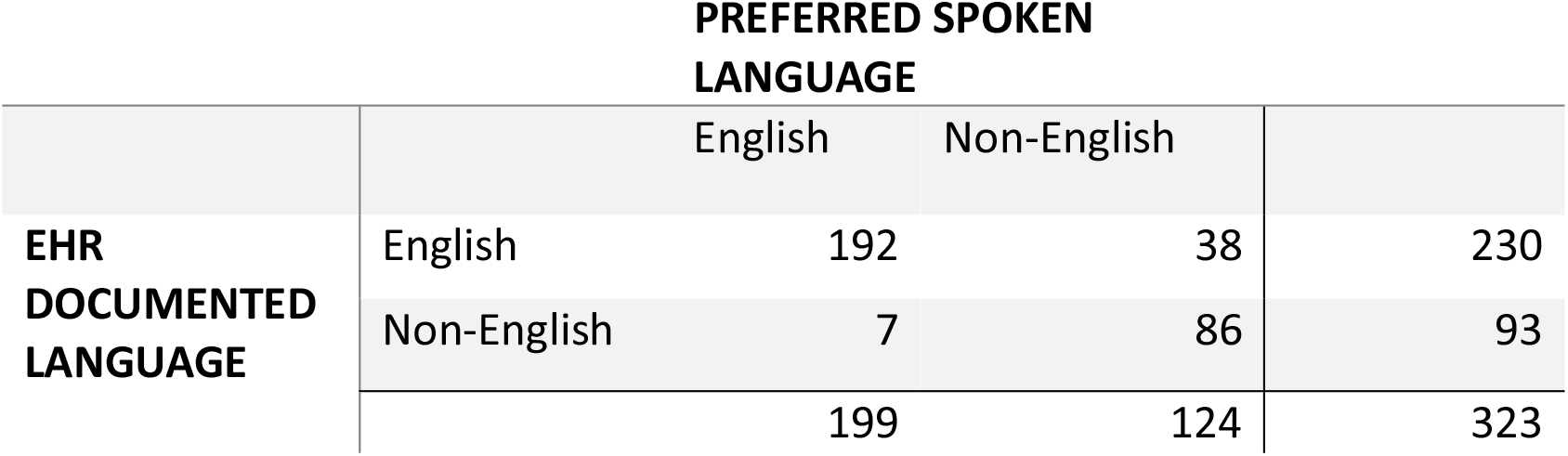
Concordance between interview- and EHR-based preferred language responses Dichotomized table (English and Non-English) of inpatient preferred spoken language data from interviews (columns) and EHR (rows). Sensitivity: 69% (CI 60 to 77). Specificity: 97% (CI 94 to 99). Positive predictive value: 95% (CI 88 to 98). Negative predictive value: 83% (CI 79 to 87). CI = 95% confidence interval. The positive condition is a non-English language preference.

The interview-based and EHR-based preferred language agreed in 278 (86%) of patients (Table 3). The probability of correctly identifying a patient with non-English preferred language (sensitivity) was 69% (CI 60 to 77); specificity was 97% (CI 94 to 99), positive predictive value was 95% (CI 88 to 98), and negative predictive value was 83% (CI 79 to 87).

**Table 3:** Languages The five most frequently recorded non-English languages (all ≥4%) during the audit. *Other languages, counts suppressed because they had fewer than 5 respondents: Albanian, Amharic, Arabic, Armenian, Dari, Farsi, Greek, Gujarati, Hakka, Hindi, Japanese, Macedonian, Mandarin, Marathi, Punjabi, Spanish, Tigrinya, Toisanese, Turkish, Urdu, Vietnamese*

| Languages Surveyed | Frequency Among Non-English Languages | Number of Respondents<sup>88</sup> |
| --- | --- | --- |
| Cantonese | 27% | 33 |
| Tamil | 14% | 17 |
| Italian | 8% | 10 |
| Tagalog | 4% | 5 |
| Bengali | 4% | 5 |
| 21 other languages | 44% | 23 |

Several factors were potentially associated with the EHR correctly identifying a non-English language preference (Figure 1). Correct identification of preferred language was less likely if a patient was male compared to female (OR 0.54, CrI 0.28 to 1.02, OR < 1 with probability 97%), cared for in Hospital 2 (OR 0.58, CrI 0.28 to 1.21, OR < 1 with probability 92%) or Hospital 3 (OR 0.68, CrI 0.35 to 1.33, OR < 1 with probability 87%) compared to Hospital 1, or cared for by the internal medicine service (OR 0.63, CrI 0.31 to 1.30, OR < 1 with probability 90%) compared to nephrology and critical care. By contrast, correct identification of preferred language was more likely for older patients (OR per decade 1.29, CrI 0.94 to 1.81, OR > 1 with probability 94%).

**Figure 1.**
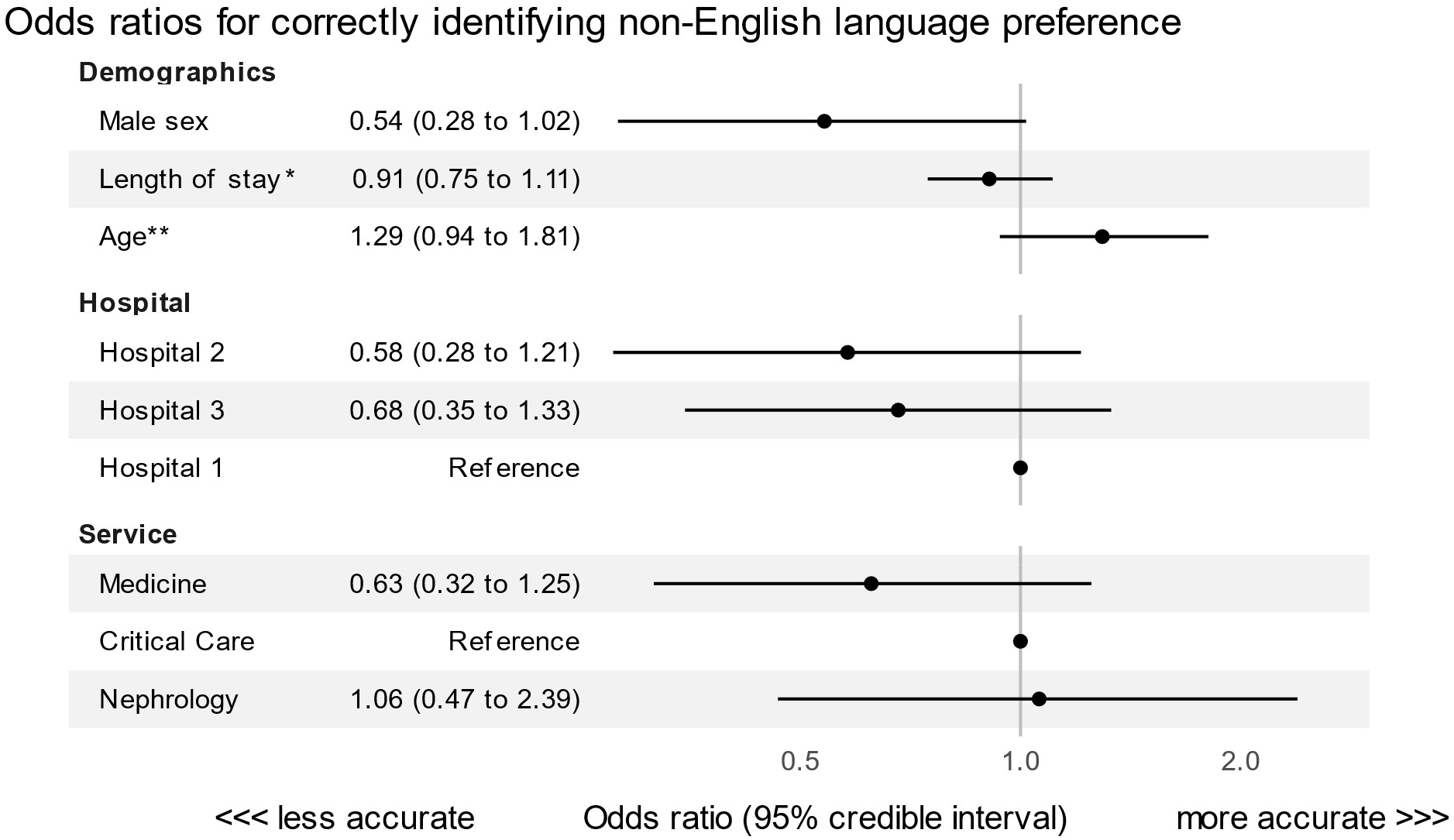
This figure shows a forest plot of odds ratios for correctly identifying EHR-based non-English language preference. This model was fit using all patients with an interview-based non-English language preference, and also included random intercepts by non-English preferred language (Figure 2). The point estimate for each odds ratio is shown as a black dot, with a horizontal black line denoting the width of the 95% credible interval. Odds ratios above 1 denote factors associated with better EHR language preference accuracy, while odds ratios below 1 denote factors associated with worse EHR language preference accuracy. * We modeled the log (base 2) of length of stay, which means that the odds ratio corresponds to each doubling of the length of stay **Age in decades was modeled as a linear predictor.

There was some variation in odds of correct identification across non-English preferred languages. The median odds ratio, which quantifies the median change in odds of correct identification when switching between two non-English preferred languages, was 1.31 (CrI 1.01 to 1.99). However, the credible intervals for the odds ratio associated with each individual language were wide (Figure 2), showing significant residual uncertainty.

**Figure 2.**
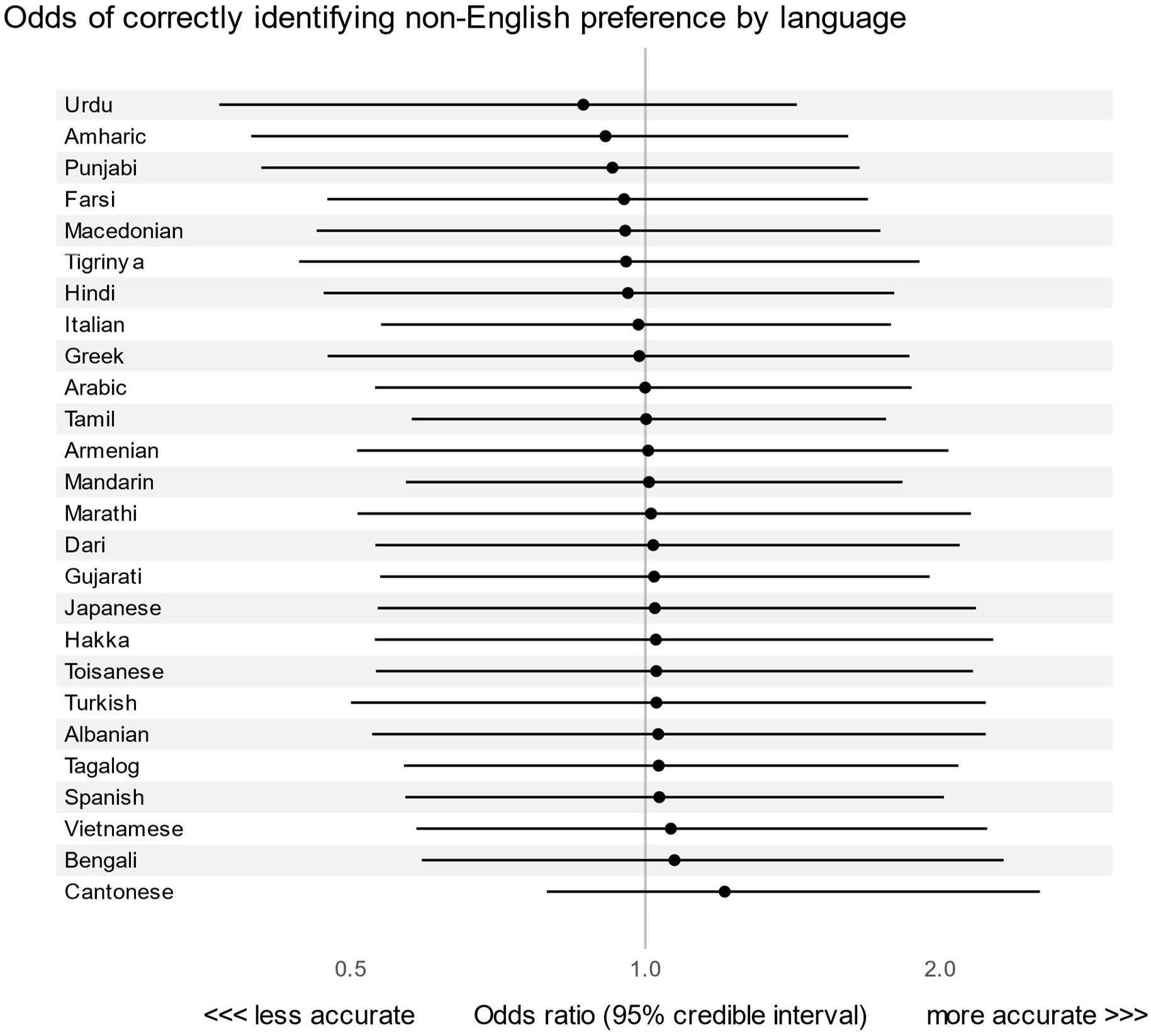
Association between non-English preferred language and the probability of the EHR correctly identifying non-English preference This figure shows a forest plot of odds ratios for EHR-based non-English language preference, showing the value of the random intercepts. This model was fit on all patients with an interview-based non-English language preference, and also included fixed predictors (Figure 1). The point estimate for each odds ratio is shown as a black dot, with a horizontal black line denoting the with of the 95% credible interval. Odds ratios above 1 denote factors associated with better EHR language preference accuracy, while odds ratios below 1 denote factors associated with worse EHR language preference accuracy. Note that no languages are convincingly associated with higher or lower probability of correctly identifying non-English preference.

## Discussion

In this prospective point-prevalence audit, we interviewed 323 patients across 3 hospitals and found that the EHR correctly captured patient preferred language in 86% of all patients, and 69% of patients who preferred a non-English language (sensitivity). The positive predictive value of a non-English preference in the EHR was 95%. Accuracy for patients who preferred non-English languages was worse for patients who were younger, and varied by hospital site and most responsible service. Non-English preferred languages comprised 26 different languages across 124 individuals. These results have implications for research investigating differences according to preferred language, highlight an opportunity to improve preferred language data, and underscore the challenges of providing language concordant care in linguistically diverse patient populations.

Prior work assessing the accuracy of EHR preferred language data had similar findings to our study. An audit in a large ambulatory care organization in California found an accuracy of 94% (15). A secondary analysis of a tobacco cessation trial in Massachusetts showed that 79% of those who preferred Spanish had that preference correctly noted in the EHR (7). A prior audit of two academic hospitals in Toronto, Canada showed sensitivities of 81% and 12% respectively (6). This last striking difference was attributed to unit clerk practices; at the hospital with higher sensitivity the clerks were trained to routinely ask about preferred language, while clerks at the other hospital only asked patients who appeared to have difficulty conversing in English. At the health network we studied, asking about preferred language is routine during patient registration for appointments or hospital admissions. Similar to past studies, our results highlight the importance of quality assurance in routinely collected data that address the social determinants of health. This is relevant for clinical care, but also relevant for operational improvements, decision support analytics, artificial intelligence, and other analyses that incorporate this data.

Our results suggest that research showing differences according to preferred language may underestimate the effect sizes of their findings.(16–18) Misclassifying patients who prefer a non-English language, which occurred in 31% of such patients in our study, would bias the findings of such studies towards the null. This means that current research may underestimate the impact of differences in care for people with non-English preferred language.

Our findings show an opportunity to improve documentation of preferred language in the EHR. The accuracy of identifying preferred language varied by hospital site and was lower overall on the medical service compared to nephrology and intensive care. The reasons for this difference are uncertain. One possible explanation for patients admitted to nephrology is that they may be more likely to have had prior interactions with the health network, allowing for more opportunities to document preferred language. The differences among hospitals could relate to differences in patient registration practices. Variation that relates to health system variables, such as responsible service or hospital site, signal opportunities to improve accurate documentation of preferred language. Potential strategies include ensuring front-line administrative staff have the permissions and knowledge to access and update preferred language, allowing patients and families to update this information themselves, and ensuring that data captured in any part of a health system is accessible to clinicians across the system.(19)

Improving accurate documentation of non-English language preference is important to enable language concordant care. Accurate measures will also enable health systems to incorporate sociodemographic variables into care delivery improvement interventions. When such measures are not considered, health systems risk widening equity-related outcome disparities.(20) For example, when patient portals were introduced to improve asthma care among low-income patients in New York, uptake was disproportionately low among high-risk elderly and non-English speaking patients.(21) Implementing new strategies to improve preferred language data collection could help ensure each patient receives language concordant care and provide a tool for incorporating equity measures into health system quality improvement activities.(22)

Among the 124 patients who preferred a non-English language, there were 26 different preferred languages. This resulted in small group sizes for each individual non-preferred language, which limited our certainty regarding language-specific variation in accuracy. In other environments there may be a single majority language among those who prefer a non-English language, such as Spanish in some US centers. Some studies evaluating outcomes by language preference specifically compare patients who prefer English to patients who prefer Spanish(23), while others study interventions designed solely for Spanish-speaking patients.(24) The same strategies or designs may not perform similarly in health care environments where there is a greater diversity of non-English preferred languages.

The need for interpretation in many different languages complicates providing language concordant care, because it limits the ability of an organization to hire full-time interpreters, complicates the development of translated non-English resources, and reduces the return-on-investment for clinicians who make efforts to learn an additional language. By contrast, telephone, video, and app-based professional interpretation services may be more helpful, due to the access to many different languages at any given time.(25,26) In the absence of professional interpreters, “ad hoc” interpreters such as family members, friends, or bilingual hospital staff often step into the medical interpretation role.(27)(28) While such interpreters have the benefit of being easily accessible, professionally trained interpreters commit fewer communication errors and preserve confidentiality.(29)(30) Collecting accurate preferred language data on an ongoing basis can help organizations decide which combinations of interpretation services will best suit their needs.

### Limitations

We did not account for bilingual or moderately fluent individuals. We did not account for individuals who may be comfortable discussing some aspects of their care in English (such as symptoms or historical details) but not others (such as advanced care planning discussions). We did not account for secondary language loss, which occurs when patients with transient or permanent neurological disease or injury revert to their first language (31). For patients who were unable to provide a response, we relied on substitute decision makers to indicate preference. Our sample size limits strong conclusions regarding accuracy according to non-English preferred language, although our analysis did reflect that uncertainty in its credible intervals.

## Conclusions

In this point-prevalence audit of a multi-hospital system, we found that the EHR accurately captured non-English language preference for 86% of all patients and 69% of patients who preferred a non-English language. Next steps will include analyzing the process of data collection and entry to improve data quality, and investigation of differences in care processes or outcomes according to preferred language.

## Data Availability

All data produced in the present study are available upon reasonable request to the authors.

## Notes

### Competing Interest Statement

The authors have declared no competing interest.

### Funding Statement

This project was supported by the Canadian Critical Care Trials Group Network of Networks Summer Student Award (CF), Sunnybrook Program to Access Research Knowledge (CF), the Nephrology Program at the Scarborough Health Network (CF), and a JP Bickell Foundation Medical Research Grant (CJY). SR is supported by an award from the Mak Pak Chiu and Mak-Soo Lai Hing Chair in General Internal Medicine, University of Toronto.

### Author Declarations

Research Ethics Board of Scarborough Health Network waived ethical approval for this work (PQI-24-002) on the basis that it was a quality improvement audit.

